# Electric field characteristics of rotating permanent magnet stimulation

**DOI:** 10.1101/2024.02.06.24302359

**Authors:** Pei L. Robins, Sergey N. Makaroff, Michael Dib, Sarah H. Lisanby, Zhi-De Deng

## Abstract

Neurostimulation devices that use rotating permanent magnets are being explored for their potential therapeutic benefits in patients with psychiatric and neurological disorders. This study aims to characterize the electric field (E-field) for ten configurations of rotating magnets using finite element analysis and phantom measurements. Various configurations were modeled, including single or multiple magnets, bipolar or multipolar magnets, rotated at 10, 13.3, and 400 Hz. E-field strengths were also measured using a hollow sphere (*r* = 9.2 cm) filled with a 0.9% sodium chloride solution and with a dipole probe. The E-field spatial distribution is determined by the magnets’ dimensions, number of poles, direction of the magnetization, and axis of rotation, while the E-field strength is determined by the magnets’ rotational frequency and magnetic field strength. The induced E-field strength on the surface of the head ranged between 0.0092 and 0.59 V/m. At the range of rotational frequencies applied, the induced E-field strengths were approximately an order or two of magnitude lower than those delivered by conventional transcranial magnetic stimulation. The impact of rotational frequency on E-field strength represents a previously unrecognized confound in clinical trials that seek to personalize stimulation frequency to individual neural oscillations and may represent a mechanism to explain some clinical trial results.

## 1. Introduction

Conventional magnetic stimulation systems, such as transcranial magnetic stimulation (TMS), utilize a current-carrying coil to generate a time-varying magnetic field pulse. This electromagnetic induction process produces a spatially varying electric field (E-field) in the brain. TMS is cleared by the United States Food and Drug Administration (FDA) for major depression, anxious depression, obsessive-compulsive disorder, smoking cessation, and migraine [1,2]. An alternative method to generating a time-varying magnetic field involves mechanically rotating permanent magnets. Several rotating magnet devices have been proposed [3–5], using high-speed rotating, high field strength neodymium magnets to induce E-field in nearby nerve tissue. The strength, spatial distribution, and precision of these E-fields in neurostimulation applications is yet to be established.

One such system, known as synchronized transcranial magnetic stimulation (sTMS) or Neuro-EEG Synchronization Therapy (NEST), has been investigated as an innovative approach to personalized treatment of major depressive disorder (MDD) [6–10]. The sTMS device consists of three cylindrical N52 grade neodymium magnets (Figure 1; Model I), diametrically magnetized with a surface field of 0.64 T [3,7,9,11]. The magnets rotate along the cylindrical axis and are positioned over the midline frontal polar brain region, the superior frontal gyrus, and the parietal region. The rotation speed of the magnets is customized to match the patient’s individual alpha frequency (IAF) of neural oscillations, as determined by pre-treatment electroencephalography (EEG) recorded from a fronto–occipital montage while the patient is in an eyes-closed resting state [9]. The hypothesized mechanism of action involves the use of exogenous subthreshold sinusoidal stimulation produced by rotating magnets to entrain alpha oscillators. This aims to reset alpha oscillators, alter neuroplasticity, normalize cerebral blood flow, and thereby improving depressive symptoms [6]. In contrast to conventional TMS, the sTMS device delivers a sinusoidal and subthreshold intensity stimulus.

**Figure 1.**
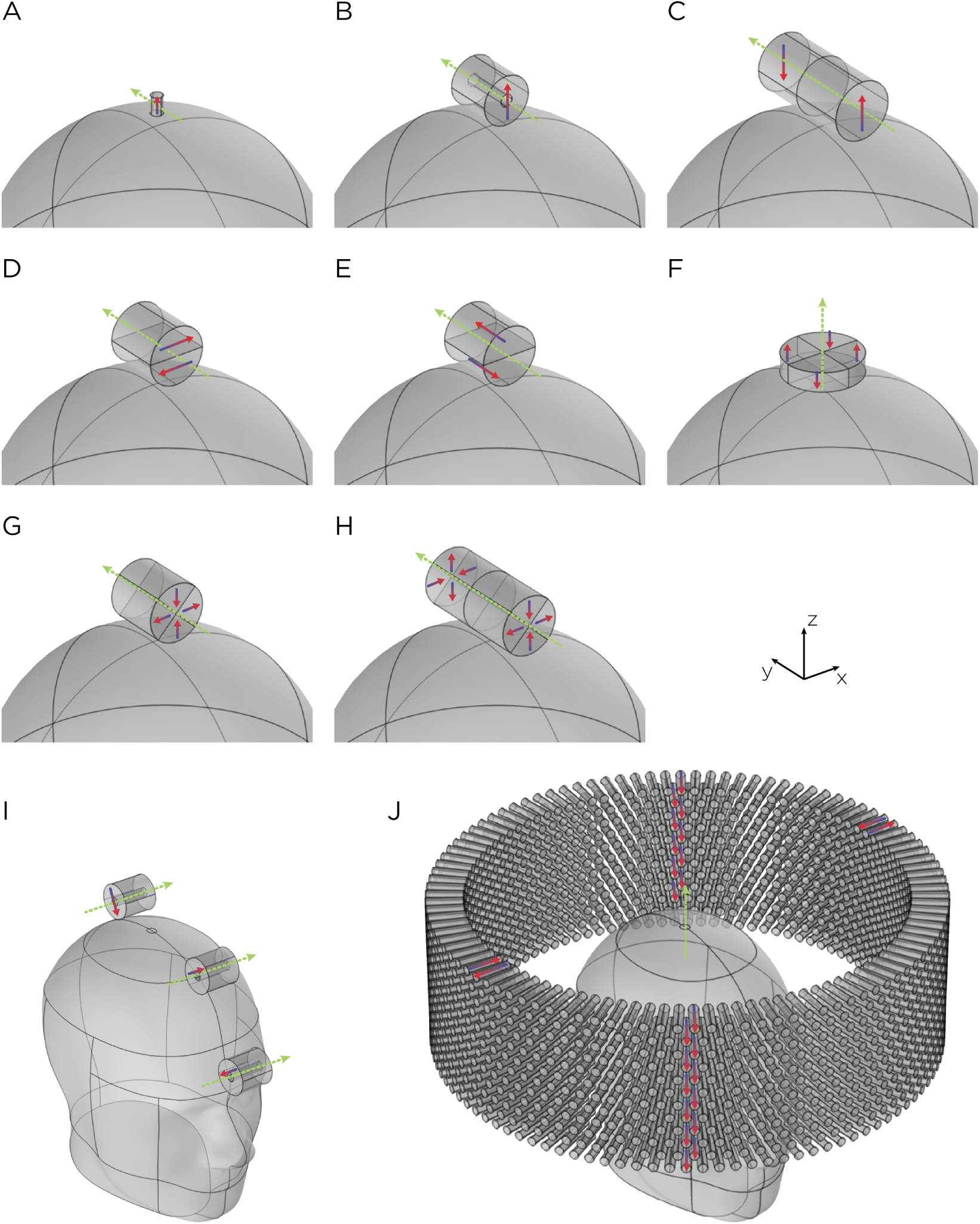
Dimensions, placement, and magnetization directions for ten configurations of rotating magnets (**A–J**). Model of single magnets in the (**A**) TRPMS and (**B**) sTMS systems. (**C**–**H**) Model of single magnets with multiple segments of different magnetization directions. (**I**) Model of the full sTMS system. (**J**) Model of the wide-bore, low-frequency magnetic spinner. The green arrows show the axes of rotation, with the direction of the rotation determined by the right-hand rule. The red/blue arrows show the direction of the magnetization.

In a multicenter, double-blinded, sham-controlled clinical trial evaluating the efficacy of sTMS for treatment of depression, no significant difference was observed between active and sham in the intent-to-treat (ITT) analysis [7]. However, among patients who completed the treatment per-protocol, there was a significant treatment response after six weeks. The authors also showed that patients in the per-protocol treatment group with a history of poor response or failed medication trials had better improvement compared to those who received no prior treatment, suggesting that more severely depressed patients may benefit more from sTMS treatment. Additionally, secondary analysis showed that lower IAF correlated with lower treatment response [8]. In addition to MDD, sTMS has also been explored as a therapeutic intervention for post-traumatic stress disorder (PTSD) [12]. In a small prospective, sham-controlled, multisite pilot of sTMS treatment for patients experiencing moderate-to-severe symptoms of PTSD, there was a greater reduction in the PTSD threshold symptoms [12]. However, there was no significant difference between the active and sham groups. Furthermore, ongoing research is assessing the safety and feasibility of sTMS in individuals with cocaine, opioid, and alcohol use disorders (ClinicalTrials.gov Identifier: NCT04336293).

Another device that employs similar mechanics is the transcranial rotating permanent magnet stimulator (TRPMS) [5,13,14]. This portable, battery-operated device consists of an array of small cylindrical N52 grade neodymium magnets mounted on high-speed motors, which are in turn mounted on a helmet. Compared to the sTMS device, the TRPMS device uses smaller magnets, measuring 0.9525 cm in height and 0.635 cm in diameter, but have stronger magnetic flux density (*B*_r_ = 1.48 T). In addition, the TRPMS magnets are axially magnetized, whereas the sTMS magnets are diametrically magnetized. However, the axis of rotation for the TRPMS magnets is perpendicular to the cylindrical axis of the magnet, whereas in the sTMS system, the axis of rotation is parallel to the cylindrical axis of the magnet. The motor operates at a no-load speed of 24 000 rpm (400 Hz), achieving a rotational speed of 20 000 rpm under load. The induced E-field strength is directly proportional to the rotational frequency of the magnet, the higher rotational speed of the TRPMS magnets results in higher E-field strength compared to the sTMS system. Voltage measurements conducted by Helekar and colleagues using an inductor search coil estimated the maximum intensity of the TRPMS device to be approximately 7% that produced by maximum conventional TMS output [14]. At a distance of 21.2 to 26.2 mm from the TRPMS and inductor, representing the depth of the cerebral cortex, the intensity reduces by approximately half.

Recent studies showed the safety and potential effectiveness of the TRPMS device in treating voiding dysfunction in patients with multiple sclerosis (MS) [15–18]. In a feasibility and safety study, the microstimulators from the TRPMS device were individually placed over predetermined regions of interest (ROI) during voiding initiation [15,17,18]. These predetermined ROIs were identified from the individual blood-oxygen-level-dependent (BOLD) activation at voiding initiation. Applying the TRPMS device to brain regions that modulate voiding initiation significantly improved bladder emptying symptoms [15,17,18]. Additionally, a proof-of-concept pilot study suggests that the TRPMS device may offer potential benefits for muscle function in individuals with type 1 myotonic dystrophy [16]. Yet another system that uses a magnet array is a wide-bore, low-frequency magnetic spinner comprised of approximately 1300 Alnico permanent magnets [19]. These magnets are arranged radially within a 30 cm diameter ring (Figure 1; Model J). The resulting rotating magnetic field is perpendicular to the ring axis, in which the measured magnetic field strength at the center of the bore is approximately 32 mT. The device reaches a rotational speed up to 15 Hz. This wide-bore magnetic spinner was originally designed to induce alternating electric currents in biological tissues, particularly in bones. Its application for brain stimulation has yet to be evaluated.

The utilization of rotating magnets has also been proposed for the stimulation of peripheral nerves and muscles [20]. Recognizing that long, straight nerves are more responsive to E-field gradients, Watterson proposed the use of multipole magnets with different magnetization directions and different axes of rotation to achieve a higher field gradient [4,20]. In several *in vitro* studies, Watterson employed a bipole magnet, featuring two diametrically magnetized cylindrical segments (N52 grade neodymium magnets with a surface field ranging from 1.43 T to 1.48 T), positioned adjacent to one another with opposite magnetization directions, to activate the sciatic nerve and attached gastrocnemius muscle in cane toad [20]. It was demonstrated that muscle and nerve activation could be achieved with rotational frequencies of 180 Hz and 230 Hz, respectively.

In this work, we assess the E-field characteristics of various rotating magnet configurations through computational modeling. Complementary to numerical simulations, experimental measurements of field strengths are performed on a head phantom, providing validation of computational results. Our objective is to provide detailed and comparative insight into the E-field profiles generated by different rotating magnet setups. We further compare their E-field characteristics to those generated by conventional TMS. Via combination of computational simulations and experimental validation, this comparative analysis aims to elucidate a comprehensive understanding of the potential advantages and limitations offered by rotating magnets for noninvasive brain stimulation applications.

## 2. Methods

### 2.1. Simulations and solver

We used finite element methods to model the rotating magnets in COMSOL Multiphysics software (COMSOL, Burlington, MA). Two different head models were used: a spherical head with a radius of 8.5 cm (Model A–H) and the Institute of Electrical and Electronics Engineers (IEEE) Specific Anthropomorphic Mannequin (SAM) phantom head (Model I–J), as illustrated in Figure 1. Both the sphere and SAM phantom head were characterized by uniform, isotropic electrical conductivity, *σ* = 0.33 S/m; relative permittivity, *ε*_r_ = 1; and relative permeability, *µ*_r_= 1. The magnets are cylindrical; they have recoil permeability, *µ*_rec_ = 1.05, which is typical of neodymium magnets [21]. The recoil permeability is the slope of the linear portion of the *B*-*H* curve, where *B* is the magnetic flux density and *H* is the magnetic field. The rotor—the moving components of the system—includes the magnet(s); the stator—the stationary part of the system—includes the head model and the surrounding air sphere.

Utilizing the magnetic vector potential (**A**–*V*) formulation and the induced solenoidal E-field, Ampère’s law was implemented across all domains. This involved the following equation:

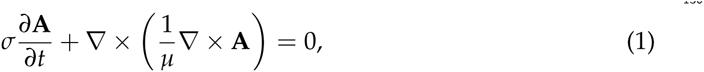

This equation signifies the relation between the time-varying component of the magnetic vector potential (**A**), the material’s conductivity (*σ*), and its permeability (*µ*). Additionally, for the sections of both the rotor and stator that were devoid of current, a magnetic flux conservation equation pertinent to the scalar magnetic potential was applied. This equation is represented as:

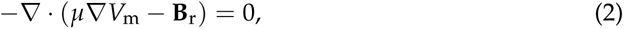

Here, *V*_m_ denotes the magnetic scalar potential, while **B**_r_ represents the remanent magnetic flux density, as detailed in [22]. Furthermore, to maintain consistency, the continuity of the scalar magnetic potential was ensured at the interface between the rotor and stator.

The stator and rotor were meshed, and then the stationary solution was computed using the multifrontal massively parallel sparse direct solver (MUMPS). Subsequently, the time-dependent aspect of the problem was addressed. This was done by solving the problem in increments of 10 degrees rotation, adhering to a relative tolerance set at 1.0 *×* 10*^−^*^8^. This approach is based on the assumption that the transient effects originating from the initiation of the rotating magnets have diminished. Consequently, the obtained final solution is indicative of the system’s steady-state behavior.

### 2.2. Multiple magnet configuration

The magnets in each model are cylindrically shaped (Figure 1). Models A and B represent single magnets from the TRPMS and sTMS systems, respectively. Model A, which measures 0.9525 cm in height and 0.635 cm in diameter, has an axial magnetization and a residual flux density of 1.48 T. This magnet is rotated about its diameter axis and tangentially to the spherical head at 400 Hz. Model B measures 2.54 cm in height and diameter, with an inner diameter of 0.635 cm. The magnet is diametrically magnetized with a residual flux density of 1.32 T and rotates about its central axis at 10 Hz.

Models C–H represent multipole configurations [4]. Model C is a bipolar magnet configuration, consisting of two diametrically magnetized cylindrical segments, each segment measures 3 cm in height and diameter, placed adjacent to each other with opposite magnetization. Model D is another bipolar configuration (3 cm in height and diameter), consisting of two diametrically magnetized, half-cylindrical segments with opposite magnetization directions. Model E (3 cm in height and diameter), similar to Model D, consists of two axially magnetized, half-cylindrical segments with opposite magnetization directions. Model F is a quadrupolar configuration (1 cm in height and 5 cm in diameter), consisting of four quadrants axially magnetized with each quadrant alternating and opposite magnetization around the central axis. The configuration is positioned on the base of the cylindrical configuration and rotates about its central axis. Model G is a quadrupolar configuration (3 cm in height and diameter), consisting of four quadrants radially magnetized with each quadrant alternating and opposite magnetization around the central axis. Model H’s configuration utilizes eight segments (6 cm in height and 3 cm in diameter), in which two Model G-like configurations are placed adjacent to each other, ensuring all eight quadrants have opposite magnetization. Configuration C–H has a residual flux density of 1.48 T and rotates about its central axis at 10 Hz.

Model I depicts the complete sTMS system, which includes three cylindrical magnets aligned along the sagittal midline of the head. The positioning of these magnets is as follows: The frontmost magnet is situated above the frontal pole, above the eyebrows; the middle magnet, positioned 7.1 cm from the frontmost magnet, aligns approximately with the superior frontal gyrus; and the most posterior magnet, located 9.2 cm from the middle magnet, corresponds roughly to the parietal cortex area. Each magnet measures 2.54 cm in both diameter and height, with an inner diameter of 0.635 cm. They are diametrically magnetized and possess a residual flux density of 1.32 T. The rotation axes are oriented perpendicular to the sagittal plane, and the rotational frequency is 10 Hz, mirroring the center frequency of the alpha band oscillation. Model J, on the other hand, represents a wide-bore, low-frequency magnetic spinner. This spinner is composed of 1224 cylindrical magnets, each 2.54 cm tall and 0.3175 cm in diameter. These magnets are axially magnetized and arranged radially within a ring with a 30 cm diameter. The magnets are uniformly distributed across 12 layers in a staggered stacking formation, with each layer being 1.905 cm apart. The spinner operates at a rotational frequency of 13.3 Hz.

### 2.3. E-field measurements

The E-field was characterized experimentally using a hollow sphere mold with a radius of 9.2 cm (Ibili, Bergara, Spain) as the head phantom, along with a custom-made silver-chloride (AgCl) twisted pair dipole probe [23]. The probe was constructed from 99.99% pure silver, 21 gauge wire, with a bare diameter of 0.635 mm, and coated with a 0.762 mm perfluoroalkoxy (PFA) layer. For insulation, the probe was coated in epoxy resin with a thickness of approximately 0.2 cm. The tips of probes are separated by a distance of 9.40 mm. The exposed tips of the probe were immersed in Clorox bleach until a light gray color was observed. The two hemispheres of the sphere mold were sealed with vacuum grease and were filled with approximately 3 liters of 0.9% sodium chloride (NaCl) in deionized water to emulate the conductivity of the brain (3.33 mS/cm at 20 *^◦^*C) [24], in which previous research have shown that 0.9% NaCl has a conductivity of 12 mS/cm at 20 *^◦^*C [25]. Figure 2 illustrates the measurement apparatus.

**Figure 2.**
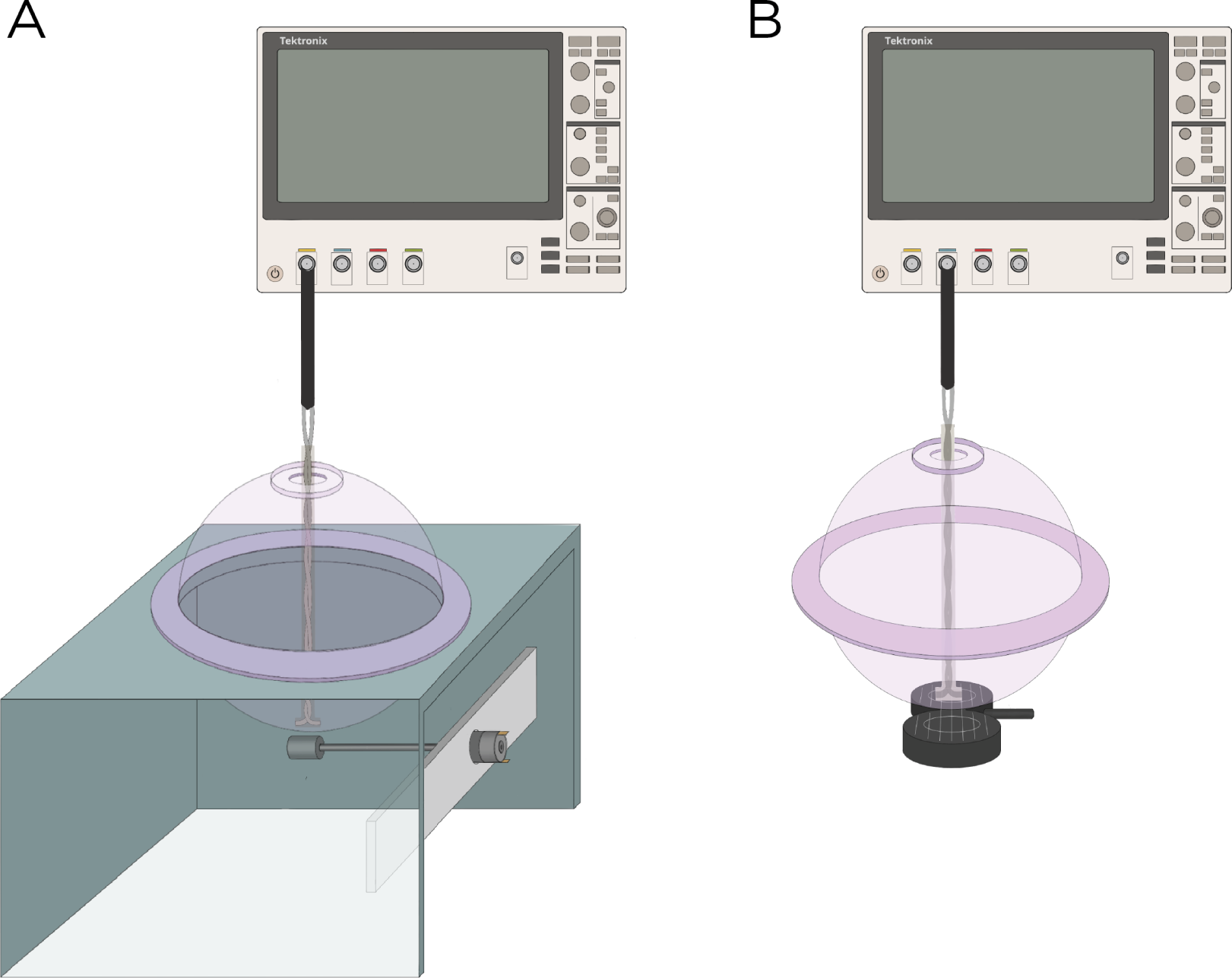
Experimental setup to measure the induced E-field strength using (**A**) Rotating magnets Models A and B and (**B**) the MagVenture TMS coil.

Model A and B were experimentally measured using magnets from K & J Magnets Inc (Figure 2A). The magnet in Model A was mounted perpendicular inside a cylinder-shaped polyetheretherketone (PEEK) material and attached to a Shengle RS550 24 V motor, enabling the magnet to rotate about its central axis and tangentially to the spherical head. The magnet in Model B had an aluminum rod attached to its inner diameter and positioned approximately 5.08 cm away from the motor to minimize interference between the magnet and the motor. Rotation of the magnet occurred along the axial direction of the cylinder. The revolution (period = *T*) of the magnets was measured using a digital hand tachometer (PH-200LC, Mitutoyo) and a piece of reflective tape (0.64 cm *×* 1.27 cm). In addition to the rotating magnets, the E-field was measured with the MagVenture TMS coil (figure-8, cooled B65 coil). The probe was oriented to measure the maximum E-field and at 100% maximum output of a MagPro X100 stimulator (Figure 2B).

## 3. Results

### 3.1. Simulations

The computational parameters and the maximum induced E-field strength for Model A–J are found in Table 1. Figure 3 illustrates the E-field distribution for Model A, representing the single rotating magnet in the TRPMS system. As the magnet rotates, the E-field distribution transitions from a figure-8 pattern (when the magnetic dipole is perpendicular to the spherical head at multiples of *T*/2) to a circular pattern (when the magnetic dipole aligns parallel to the head at multiples of *T*/4). The peak induced E-field strength at the surface of the head is approximately 0.59 V/m, in the direction parallel to the rotation axis of the magnet. Figure 4, representing the single magnet in the sTMS system (Model B), presents a similar E-field distribution as Figure 3 at a lower E-field strength. The peak induced E-field strength at the head’s surface for this magnet configuration measures approximately 0.098 V/m in the direction perpendicular to the direction magnetization.

**Figure 3.**
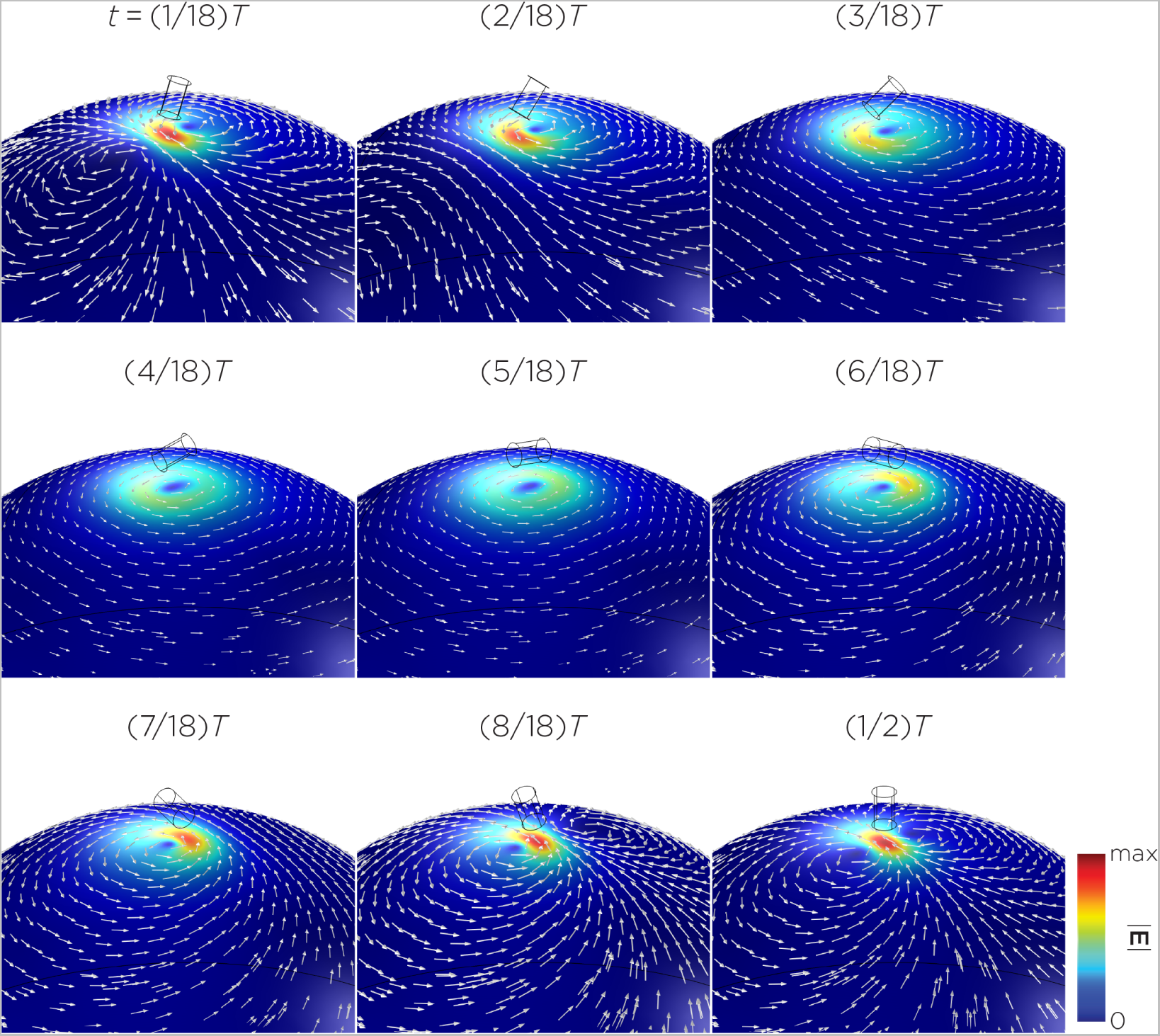
Half revolution of configuration A in steady-state.

**Figure 4.**
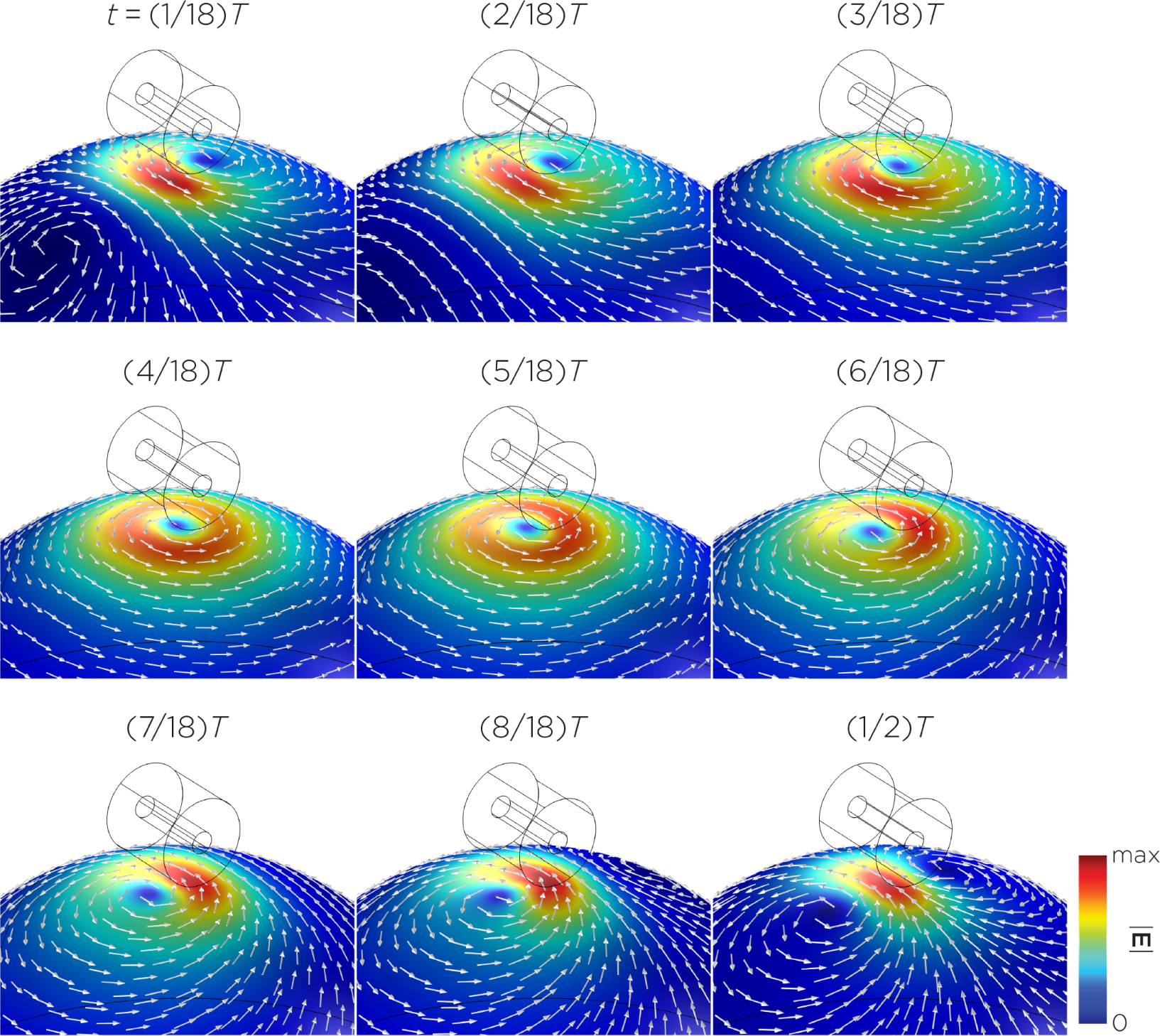
Half revolution of configuration B in steady-state.

**Table 1.** Table caption.

| Model | Dimensions<br>(cm) | Magnetization<br>direction | $B_r$<br>(T) | Angular<br>frequency (Hz) | max $\vert \mathbf{B}\vert $<br>(mT) | max $\vert \mathbf{E}\vert $<br>(V/m) |
| --- | --- | --- | --- | --- | --- | --- |
| A | Cylinder × 1<br>od = 0.635<br>h = 0.9525 | Axial | 1.48 | 400 | 94.1 | 0.59 |
| B | Ring × 1<br>od = 2.54<br>id = 0.635<br>h = 2.54 | Diametrical | 1.32 | 10 | 334.8 | 0.098 |
| C | Cylinder × 1<br>2 segments<br>od = 3, h = 6 | Diametrical,<br>multipole | 1.48 | 10 | 462.8 | 0.13 |
| D | Cylinder × 1<br>2 segments<br>od = 3, h = 3 | Diametrical,<br>multipole | 1.48 | 10 | 209.8 | 0.13 |
| E | Cylinder × 1<br>2 segments<br>od = 3, h = 3 | Axial,<br>multipole | 1.48 | 10 | 134.1 | 0.025 |
| F | Cylinder × 1<br>4 segments<br>od = 5, h = 1 | Axial,<br>multipole | 1.48 | 10 | 260.7 | 0.13 |
| G | Cylinder × 1<br>4 segments<br>od = 3, h = 3 | Radial,<br>multipole | 1.48 | 10 | 353.7 | 0.23 |
| H | Cylinder × 1<br>8 segments<br>od = 3, h = 6 | Radial,<br>multipole | 1.48 | 10 | 350.6 | 0.14 |
| I | Ring × 3<br>od = 2.54<br>id = 0.635<br>h = 2.54 | Diametrical | 1.32 | 10 | 354.7 | 0.11 |
| J | Cylinder × 1224<br>od = 0.3175<br>h = 2.54<br>Array id = 30<br>12 layers<br>layer separation = 1.905 | Axial | 1.48 | 13.3 | 2.5 | 0.0092 |

h: height; id: inner diameter; od: outer diameter

Figure 5 displays a bipolar E-field distribution in Model C. As the magnet rotates, the E-field distribution shifts from a four-leaf clover pattern (when the magnetization direction is perpendicular to the spherical head at multiples of *T*/2) to a figure-8 pattern (at multiples of *T*/4). The peak induced E-field strength at the head’s surface measures approximately 0.13 V/m. Figure 6 (Model D) showcases another bipolar E-field distribution similar to Models A and B. Similarly, the circular pattern occurs when the magnetization directions are parallel to the spherical head. In this configuration, the peak induced E-field strength measures approximately 0.13 V/m. Additionally, Figure 7 (Model E) shows a bipolar E-field distribution with a similar pattern as Figure 5 (Model C), with lower peak induced E-field strength of approximately 0.025 V/m. Figure 8 (Model F) demonstrates a quadrupolar E-field distribution. As the magnet rotates, the E-field distribution is a four-leaf clover that rotates. The peak induced E-field strength measures approximately 0.13 V/m. Figure 9 is another quadrupole E-field distribution, with similar E-field patterns as Models A, B, and D. In this configuration, the peak induced E-field strength is approximately 0.23 V/m.

**Figure 5.**
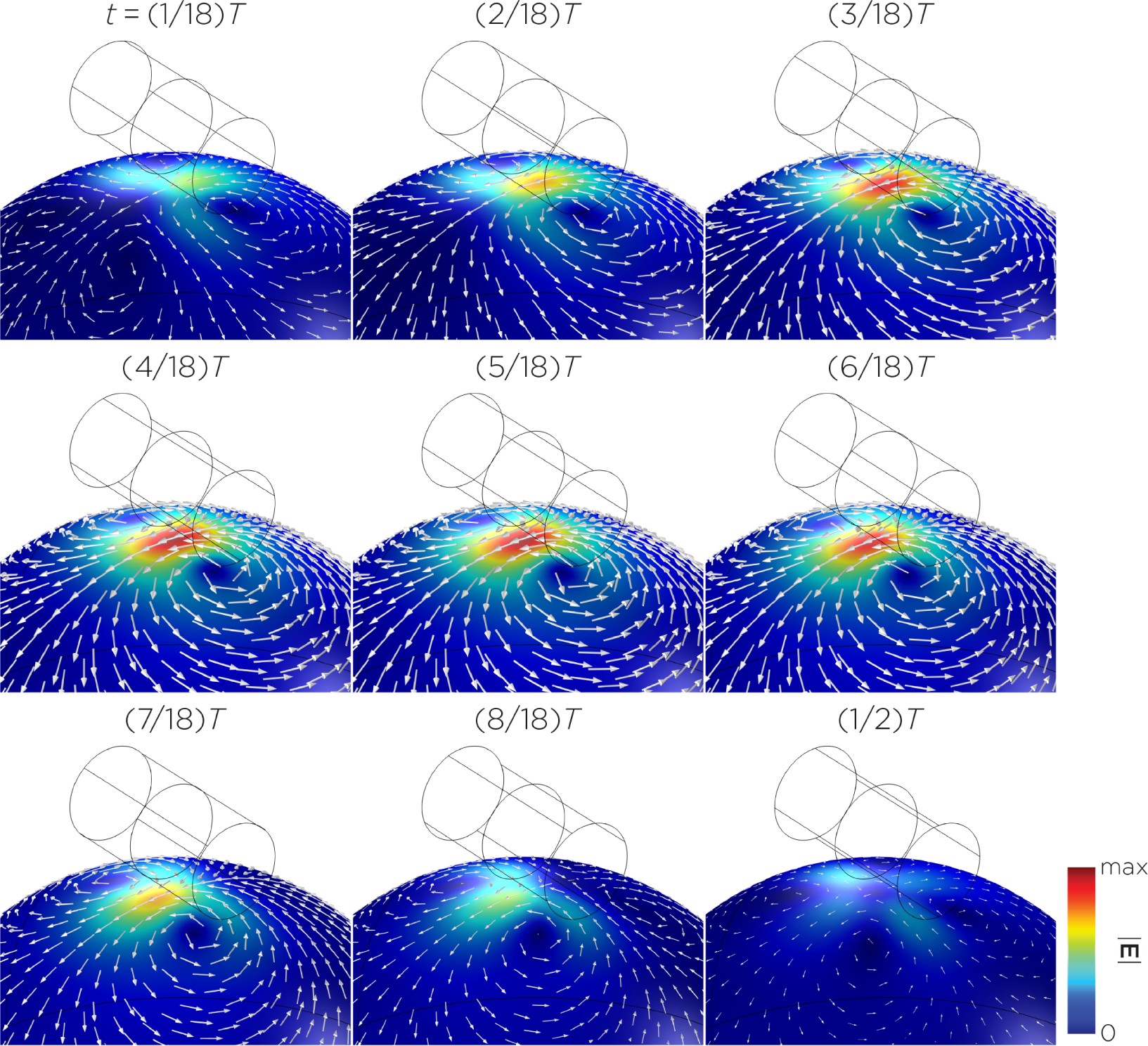
Half revolution of configuration C in steady-state.

**Figure 6.**
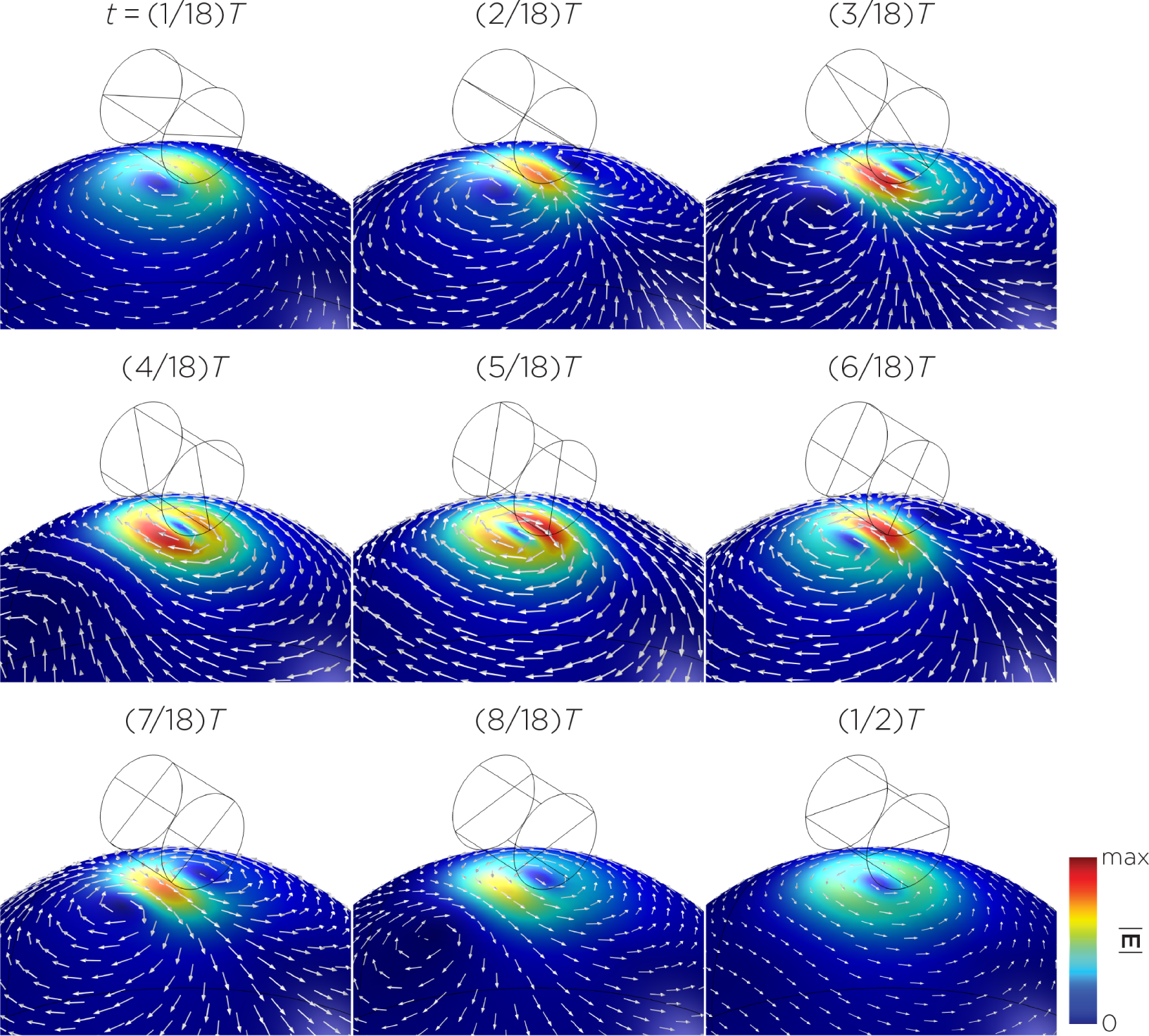
Half revolution of configuration D in steady-state.

**Figure 7.**
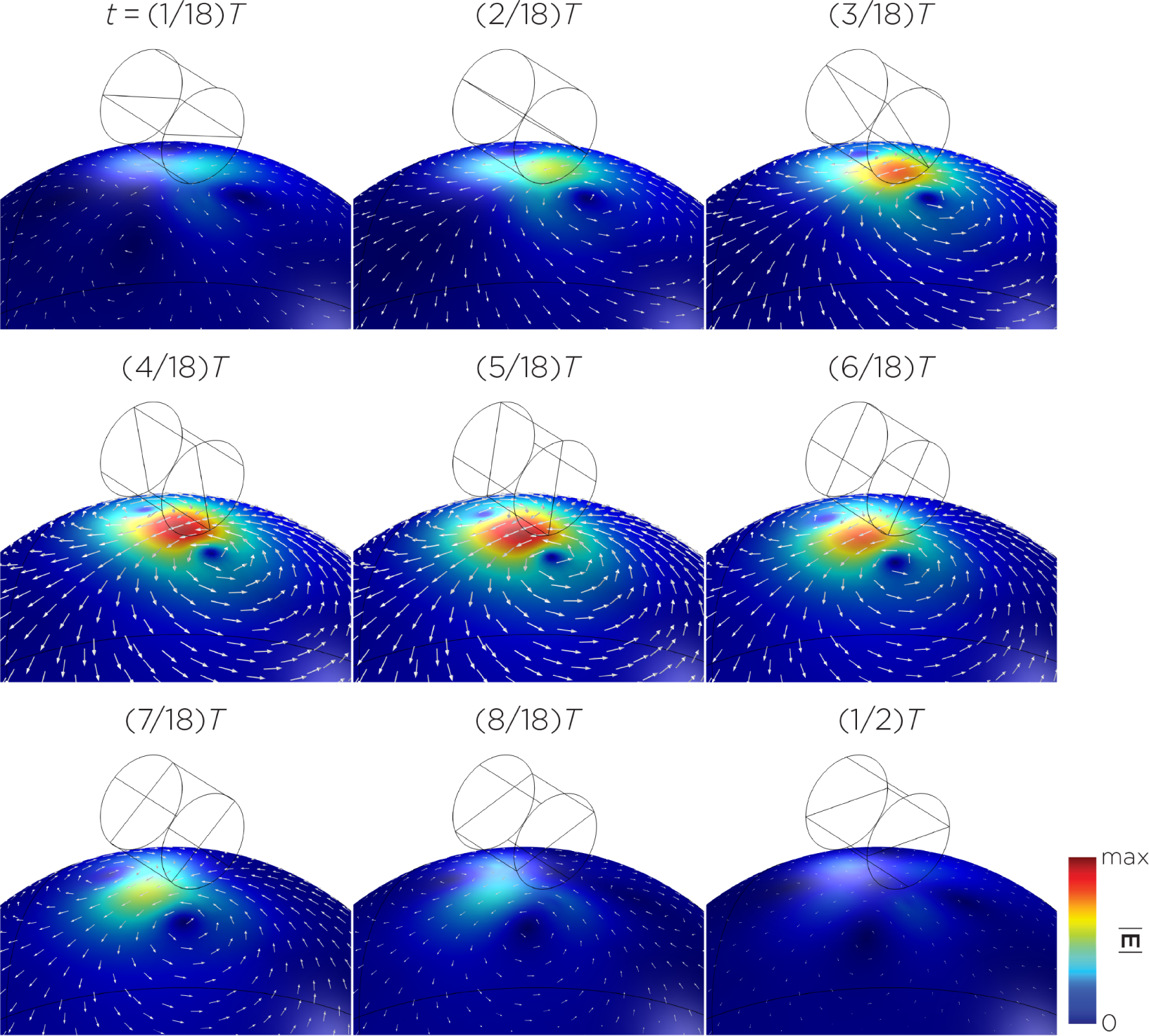
Half revolution of configuration E in steady-state.

**Figure 8.**
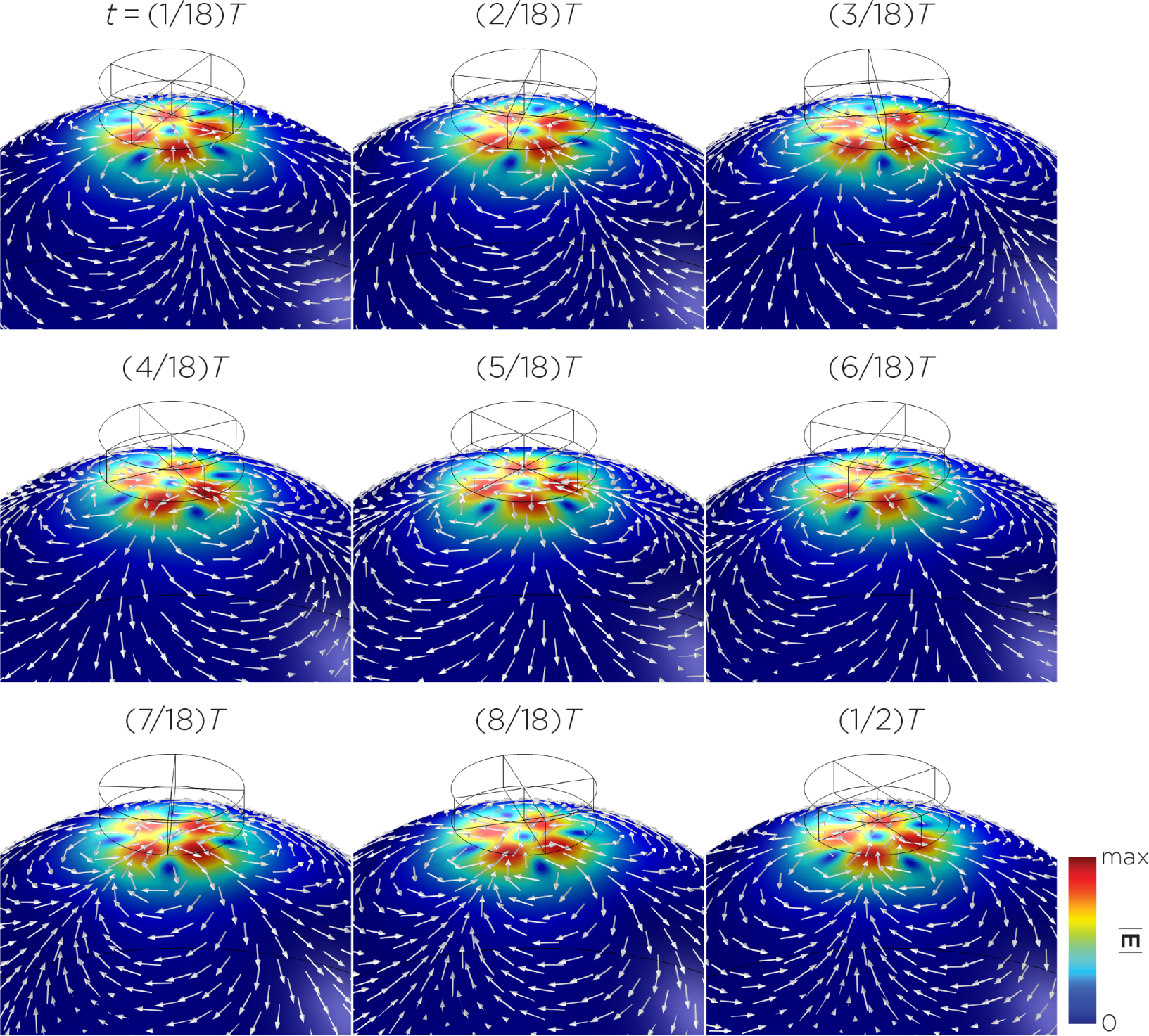
Half revolution of configuration F in steady-state.

**Figure 9.**
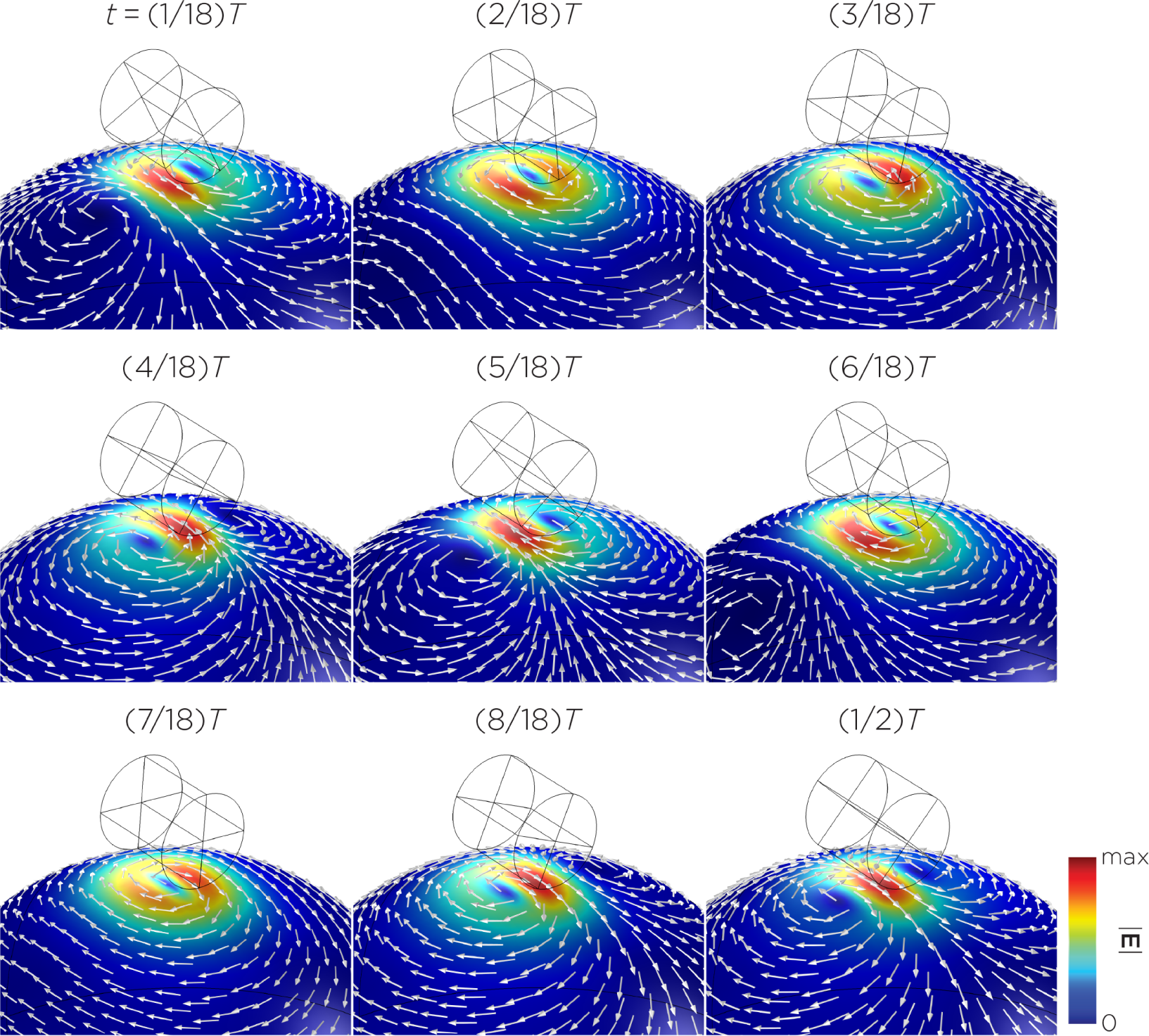
Half revolution of configuration G in steady-state.

Figure 10 shows an 8-pole E-field distribution with a similar E-field distribution as Model C. The peak induced E-field strength measures approximately 0.14 V/m.

**Figure 10.**
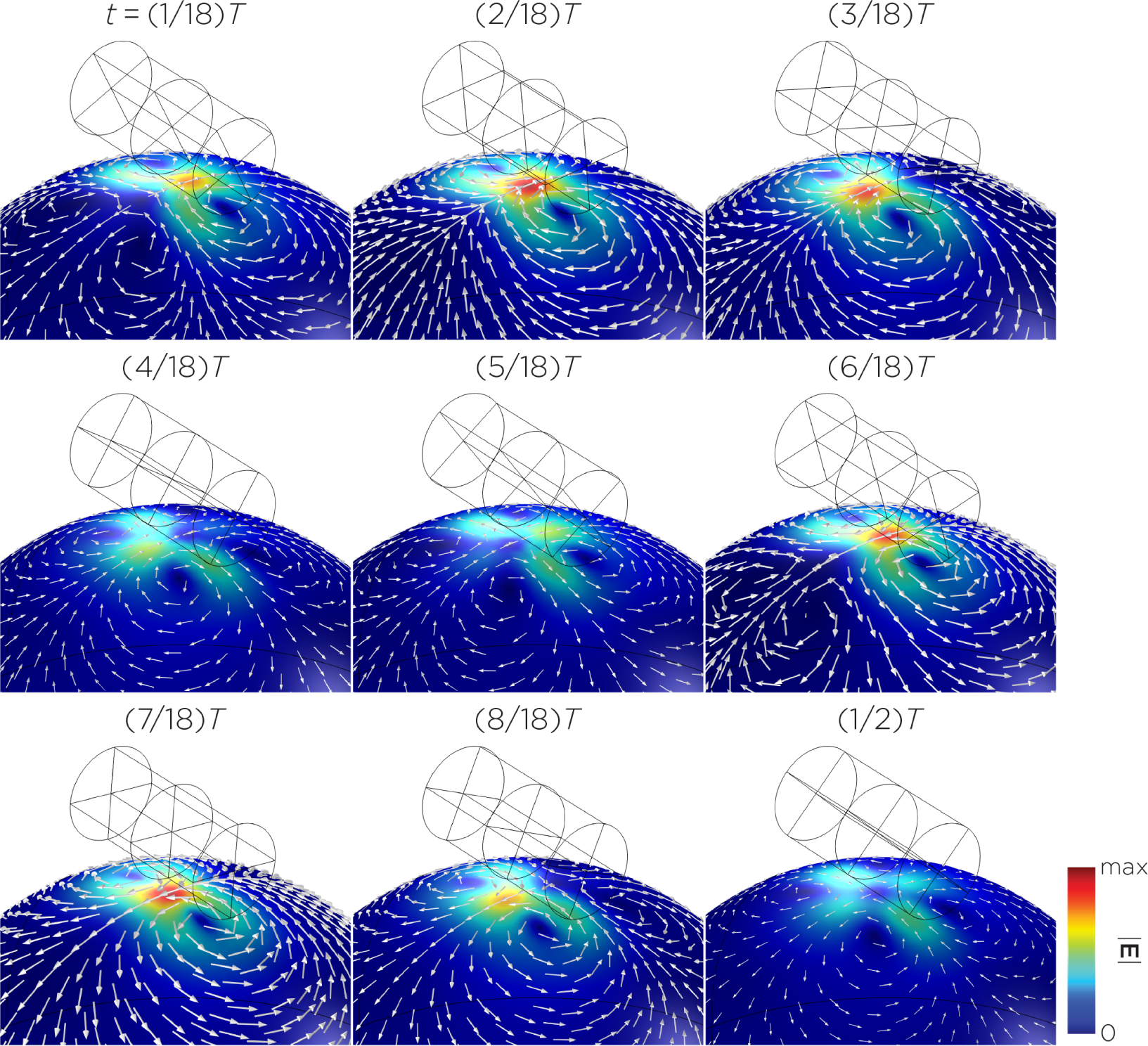
Half revolution of configuration H in steady-state.

Figure,11 illustrates the E-field distribution for the three-magnet sTMS system. In this configuration, the stimulation is extensively spread across the midline frontal polar, medial frontal, and parietal regions of the head. The maximum E-field strength induced at the head’s surface is approximately 0.11 V/m. Notably, at a depth of 1.5 cm from the surface of the head, which is roughly the depth of the cortex, the strength of the E-field reduces by about half. Figure,12 displays the E-field distribution generated by the widebore, low-frequency magnetic spinner. Here, the stimulation is broadly distributed in a vertical orientation relative to the head and rotates around the head as the spinner device rotates. The peak E-field strength induced at the surface of the head in this configuration is approximately 0.0092 V/m.

**Figure 11.**
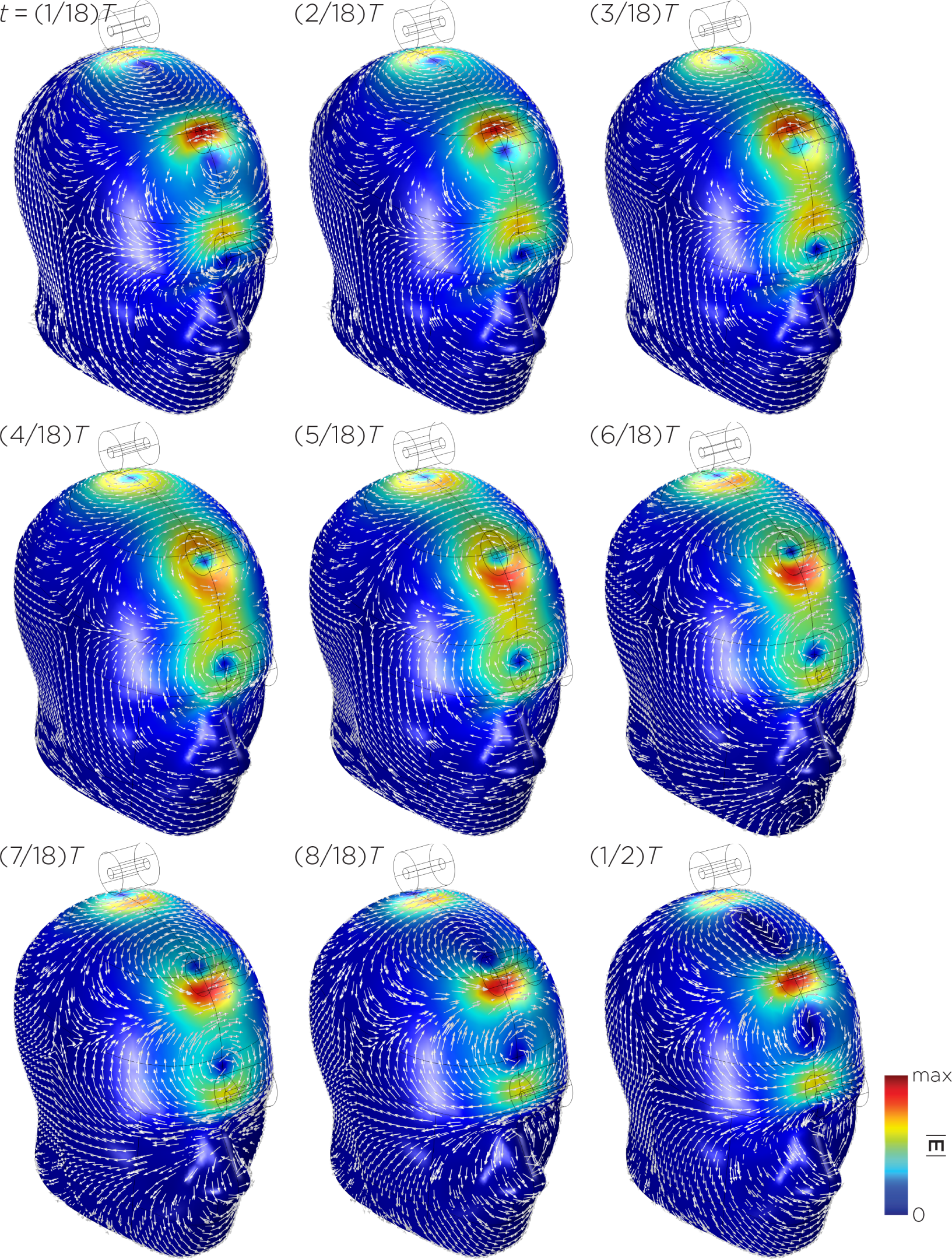
Half revolution of configuration I in steady-state.

**Figure 12.**
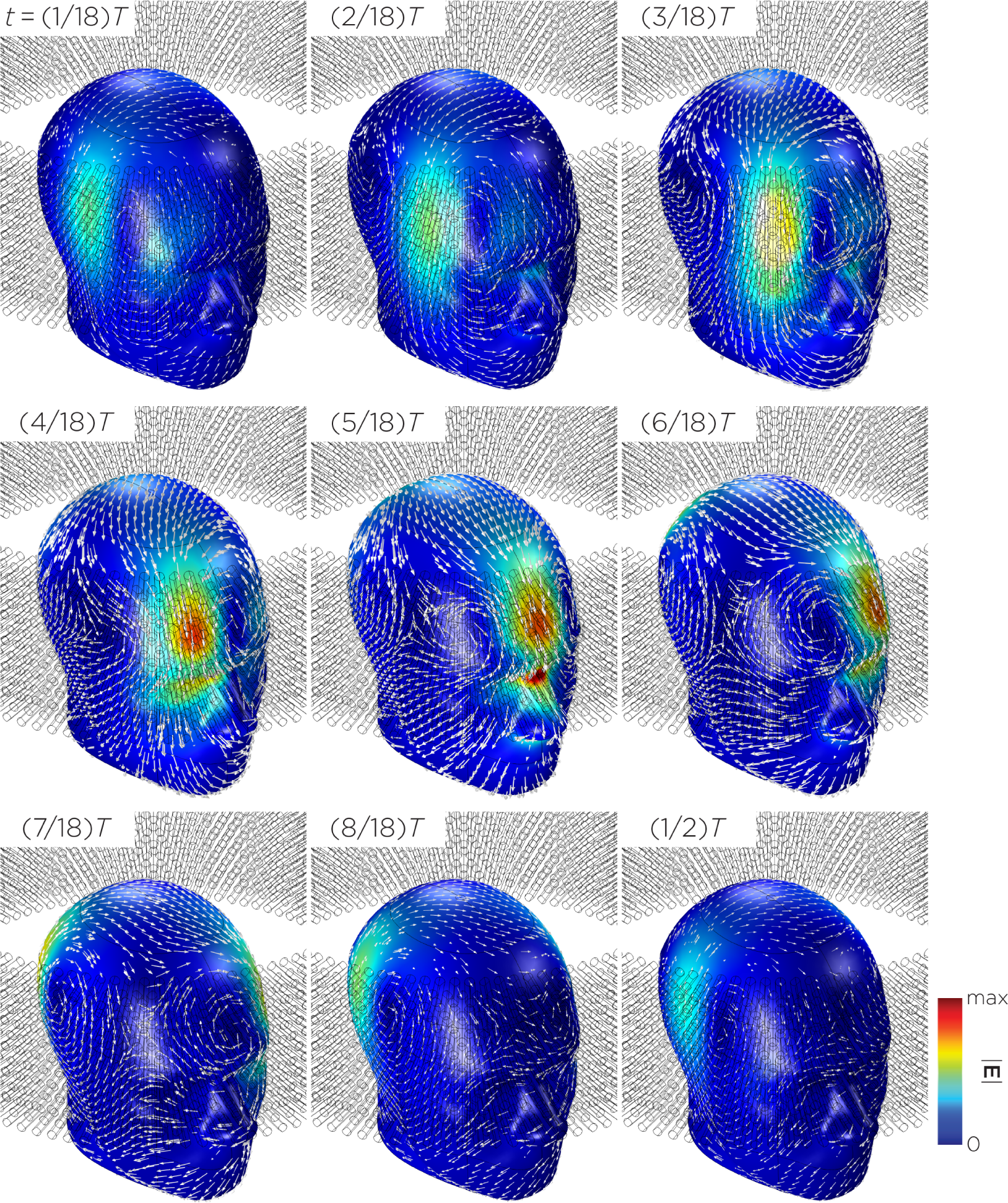
Half revolution of configuration J in steady-state.

### 3.2. Experimental measurements

Model B, representing one single magnet of the sTMS system, induces a maximum Efield strength (0.098 V/m) lower compared to the maximum induced E-field strength of the sTMS system (0.11 V/m). When comparing the E-field measurements to the computational results for Model A and B, there are similar values reported in Table 2. Specifically, the maximum E-field for Model A was found to be approximately 0.39 V/m when the magnets were spun at 349.9 Hz and 0.082 V/m at 10.1 Hz for Model B. The maximum E-field for the MagVenture TMS coil was measured to be approximately 401.5 V/m at a pulse frequency of 3448 Hz.

**Table 2.** E-field measurements.

| Configuration | Rotational/Pulse frequency (Hz) | max $\vert E\vert $ (V/m) |
| --- | --- | --- |
| Model A | 349.9 | 0.39 |
| Model B | 10.1 | 0.082 |
| TMS | 3448 | 401.5 |

When comparing the E-field measurements to the computational results for Model A and B, there are similar values reported in Table 2. Specifically, the maximum E-field for Model A was found to be approximately 0.39 V/m when the magnets were spun at 349.9 Hz and 0.082 V/m at 10.1 Hz for Model B. The maximum E-field for the MagVenture TMS coil was measured to be approximately 401.5 V/m at a pulse frequency of 3448 Hz. Model B, representing one single magnet of the sTMS system, induces a maximum E-field strength (0.098 V/m) comparable to the maximum induced E-field strength of the full sTMS system (0.11 V/m).

## 4. Discussion

The distribution of the E-field induced by a single rotating magnet is influenced by several factors, including the dimensions and placement of the magnet(s), the number of poles, direction of the magnetization, and axis of rotation. Furthermore, the E-field strength is dependent on the rotational frequency and the surface field strength of the magnets. Our simulations revealed that the induced E-field strength on the head surface ranged from 0.0092 V/m to 0.59 V/m. The spatial pattern of the E-field varied between circular, figure-8, or four-leaf clover shapes, depending on the relative orientation of the magnetization vector to the head model. For instance, in Model A, the E-field exhibits a circular pattern when the magnetization vector is parallel to the head and morphs into a figure-8 pattern when the magnetization vector is rotated perpendicular to the head. With more than one magnet, the E-fields from each magnet are summated according to the principle of superposition, resulting in more complex patterns and strengths depending on the arrangement and characteristics of the magnets (e.g., the distance between the magnets and their initial polarizations). Similarly, single magnet configurations with multiple poles illustrated complex patterns and strengths, depending on the summated magnetization directions. In general, configurations with a figure-8 or more localized E-field distribution resulted in higher peak surface E-field strength, while those with a circular or more spreadout pattern induced lower peak surface E-field strength. This phenomenon mirrors the depth–focality trade-off observed in TMS coils, where the E-field strength in a more focal distribution decays more rapidly with distance compared to a more spread-out E-field distribution [26].

In Model I of the full sTMS device, the rotational frequency of the magnets were set to 10 Hz. Given that the strength of the induced E-field is directly proportional to the rotational frequency, it becomes feasible to calculate the field strengths at different frequencies. In practical applications, the sTMS system aligns the rotational frequency of the magnets with the participant’s IAF, as measured by EEG, which typically falls within the 8 to 13 Hz range [9]. Jin and Phillips (2014) approximated the intensity of sTMS to be less than 1% of that produced by conventional TMS. This estimation was based on comparing the maximum rate of change in the magnetic field over time (d*B*/d*t*) between the sinusoidal waveform used in sTMS and the pulsed waveform of conventional TMS. However, this comparison overlooked critical factors such as the spatial characteristics of the magnetic fields and the boundary conditions of the head, which significantly influence the distribution and intensity of the induced E-field. Our simulations and measurements with a single magnet (Model B), as well as simulations of the complete three-magnet array (Model I), have identified a peak E-field strength of approximately 0.1 V/m. This value signifies that the strength at the surface of the head is only about 0.025% of that generated by conventional TMS, indicating a considerably lower intensity in comparison.

Synchronizing the exogenous subthreshold sinusoidal stimulation to the intrinsic alpha EEG rhythm was thought to be an important feature that underlies the mechanism of sTMS treatment for depression [6]. In the sTMS depression study, some participants did not receive stimulation at the correct IAF, which led to inferior outcomes compared to those treated at the correct IAF [7]. Secondary analysis showed that participants with lower IAF exhibited the least clinical improvement [8]. It is important to note that since the induced E-field strength is directly proportional to the rotational frequency of the magnets, customizing the rotational frequency could introduce variability in the induced E-field strength across individuals. In addition, a lower IAF may not only represent a trait of treatment resistance but would also determine a lower rotational frequency of the magnets, which would in turn lead to a decreased E-field strength, thereby potentially confounding the interpretation of this finding. One potential solution we propose to mitigate this confound is to employ electromagnets, such as those used in n-phase motors. In a 3-phase motor, for example, three coil windings in the motor stator receive power from three alternating currents that are out of phase with each other by one-third of their cycle, creating a magnetic field that rotates, similar to that in a mechanically rotating permanent magnet system. The advantage of using electromagnets is that it independently controls frequency and amplitude, as opposed to the fixed coupling of these variables in mechanical rotating systems.

Compared to the sTMS system, the TRPMS system produces slightly higher E-field strength, using a stronger magnet and a much higher rotational frequency. Inductive measurements by Helekar and colleagues estimated the maximum TRPMS stimulation intensity to be approximately 7% of maximum conventional TMS output at a distance of 6.2 mm from the magnet or TMS coil [14]. First, these estimates are based on measurements made in the air and do not account for head boundary conditions, potentially overestimating the E-field strength. Second, the TMS waveform reported in Helekar et al. [14] does not resemble the conventional biphasic cosine waveform generated by the Magstim Rapid^2^ stimulator. This is possibly due to a lower sampling rate in their measurements, causing a distortion in the waveform, underestimating the peak value. Third, since smaller magnets have faster field attenuation with distance compared to larger magnets [26], the E-field strength of the TRPMS system at a depth of the cortex would be further overestimated. Our simulation and measurement for a single magnet in the TRPMS system showed that the peak E-field strength is approximately 0.1% of conventional TMS.

In the simulations of multipole magnets rotating at 10 Hz (Model C–H), the E-field strengths are similar to that of the sTMS system, except for Model E, which exhibits an E-field strength (0.025 V/m), approximately an order of magnitude lower. The E-field distribution from Watterson’s configurations demonstrated characteristics of multiple magnets. For example, Model C, representing the bipole configuration used in Watterson’s nerve stimulation experiments, exhibits a four-clover and a figure-8 shaped field pattern. The four-clover pattern emerges when the magnetization direction is perpendicular and shifts to a figure-8 pattern when the magnetization direction is parallel to the head model. Using this bipole configuration, Watterson and colleagues demonstrated the ability to achieve nerve activation at a rotational frequency of 230 Hz [20]. According to their measurements, this resulted in an E-field strength of approximately 1 V/m, equivalent to 0.4% of maximum conventional TMS output. In our finite element models, we use a rotational frequency of 10 Hz to simulate the effect of multipolar magnet stimulation for brain stimulation. The 10 Hz frequency matches that of the sTMS model. Our simulation shows that this configuration induces an E-field strength of 0.13 V/m, approximately 0.032% of conventional TMS at the surface of the head. The multipolar magnets could be as effective as sTMS when used as part of a brain stimulation device.

In comparison to other proposed rotating magnetic systems, the wide-bore, low-frequency magnetic spinner (Model J), designed to induce alternating electric currents in biological tissues, induced the lowest and most nonfocal E-fields. This device generates a maximum magnetic field of 2.5 mT, resulting in a maximum induced E-field of 0.0092 V/m in the head model. With the installation of a magnetic yoke, which concentrates the magnetic flux to the inside of the bore, the measured magnetic field reaches 32 mT, bringing the induced E-field strength close to that of other devices. In terms of the spatial distribution, there are two E-field peaks located where the column of magnets reverses magnetization, i.e., where the magnetic field gradient is the highest. Since the induced field is more diffused, the field penetration is deeper compared to other smaller rotating magnetic configurations.

One potential advantage of utilizing rotating permanent magnets is the ability to create portable, cost-effective devices compared to conventional TMS [20]. Depending on the magnet strength and rotational frequency, the E-field strengths in the sTMS, TRPMS, and Watterson multipolar systems are comparable to other forms of low field stimulation, including low field magnetic stimulation (LFMS) [27], transcranial current stimulation (tCS) [28,29], and low-intensity repetitive magnetic stimulation (LI-rMS) [30,31]. Low field stimulation has been shown to induce changes at the cellular and molecular levels. For example, in an *in vitro* model, LI-rMS has demonstrated the capability to modify cellular activation and gene expression patterns in neural tissue explants [30]. This modulation is distinctive and dependent on the specific frequencies and pattern of stimulation employed. Dufor and colleagues reported induced E-field strengths of this device to be between 0.05 V/m to 0.075 V/m [32]. Similarly, LI-rMS delivered during visually-evoked activity increased densities of parvalbumin-expressing GABAergic interneurons in adult mouse visual cortex [33]. These findings suggest that the low field strengths produced by rotating permanent magnets might be biologically active, warranting further investigation to evaluate their potential therapeutic value.

Achieving higher field strengths through increased rotational speeds of the magnets is feasible. However, it’s crucial to consider the low-pass filtering characteristic of the neuronal membrane. Due to this property, rapid voltage changes are not transmitted as efficiently across the membrane, which can reduce the effectiveness of high-frequency stimulation [34]. Moreover, the relationship between field strength and excitation frequency may be nonlinear. This nonlinearity has been evidenced in studies involving transcranial alternating current stimulation (tACS), such as those conducted by Moliadze et al. (2012) and Numssen et al. (2021) [35,36]. For instance, in Moliadze et al. (2012)’s study [35], when 140 Hz tACS was applied to the motor cortex, a low current amplitude of 0.4 mA led to a decrease in motor evoked potential (MEP) amplitudes. Conversely, intermediate amplitudes of 0.6 mA and 0.8 mA showed no significant effect on MEPs. At a higher amplitude of 1 mA, there was an enhancement of MEP amplitudes. This example underscores the complexity of interactions between field strength and stimulation frequency, suggesting that adjustments in stimulation parameters may yield varying and potentially unexpected effects on neural responses.

Several limitations should be acknowledged when interpreting this work. First, we did not perform a high-resolution spatial sampling of the E-field in the phantom to characterize the full spatial distribution, which varies over time. Our focus was on measuring the peak E-field strength to validate the simulation magnitudes and compare them to previously conducted measurements. Second, the simulations were not performed on realistic head models. Our models assume a simplified geometry with homogeneous, isotropic conductivity to better illustrate the spatial field distribution. Cortical folding in realistic models can increase the maximum E-field strength compared to spherical head models [28,37]. Future work could consider integrating realistic head models to better represent actual head anatomy and the variations in E-field strengths across individuals.

## 5. Conclusions

In our study, we performed a thorough examination of the E-field in rotating magnet systems, including the sTMS device, the TRPMS device, and various other configurations of rotating magnets. This was achieved through computational modeling and E-field measurements using a phantom head. Our results revealed that the maximum E-field strength induced at the head’s surface varied between 0.0092 V/m to 0.59 V/m. This range corresponds to approximately 0.1% of the field strength generated by conventional TMS. Notably, our investigation highlights the impact of the rotational frequency of the magnets on E-field strength. This finding introduces a previously unappreciated confound in clinical trials that aim to synchronize the rotational frequency with individual endogenous oscillatory activities. Looking forward, our future research endeavors include conducting simulations of rotating magnetic stimulation on anatomically realistic head models derived from individual brain imaging data. We also plan to optimize treatment parameters, such as stimulation frequency and magnet placement, with the aim of enhancing therapeutic efficacy. Additionally, obtaining direct electrophysiological data is crucial to validate the proposed mechanism of action of these stimulation systems. Such data would be instrumental in understanding the physiological effects of these technologies, ultimately allowing for refinement of their clinical potential.

## Author Contributions

Conceptualization, Z.-D.D. and P.L.R.; methodology, P.L.R., S.N.M., and Z.-D.D.; validation, P.L.R. and Z.-D.D.; formal analysis, P.L.R. and Z.-D.D.; data curation, P.L.R. and Z.-D.D.; writing—original draft preparation, P.L.R. and Z.-D.D.; writing—review and editing, P.L.R., S.N.M., M.D., S.H.L., and Z.-D.D.; visualization, P.L.R. and Z.-D.D.; supervision, Z.-D.D. All authors have read and agreed to the published version of the manuscript.

## Data Availability Statement

The raw data supporting the conclusions of this article will be made available by the authors on request.

## Acknowledgments

This work utilized the computational resources of the NIH HPC Biowulf cluster (http://hpc.nih.gov). The authors also thank the NIMH Section on Instrumentation for helping to build the measurement apparatus. The opinions expressed in this article are the author’s own and do not reflect the views of the National Institutes of Health, the Department of Health and Human Services, or the United States government.

